# The influence of training load on hematological Athlete Biological Passport variables in elite cyclists

**DOI:** 10.1101/2020.10.22.20213413

**Authors:** Astolfi Tiffany, Crettaz von Roten Fabienne, Kayser Bengt, Saugy Martial, Faiss Raphael

**Affiliations:** REDs, Research and Expertise in antiDoping sciences, University of Lausanne, Switzerland; ISSUL, Institute of Sport Sciences, University of Lausanne, Switzerland

**Author notes:** Correspondence: Raphael Faiss.

**Keywords:** blood, training load, hemoglobin, plasma volume, anti-doping

## Abstract

The hematological module of the Athlete Biological Passport (ABP) is used in elite sport for antidoping purposes. Its aim is to better target athletes for testing and to indirectly detect blood doping. The ABP allows to monitor hematological variations in athletes using selected primary blood biomarkers (hemoglobin concentration ([Hb] and reticulocyte percentage (Ret%)) with an adaptive Bayesian model to set individual upper and lower limits. If values fall without the individual limits, an athlete may be further targeted and ultimately sanctioned.

Since [Hb] and Ret% vary with plasma volume (PV) fluctuations, possibly caused by training load changes, we investigated the putative influence of acute and chronic training load changes on the ABP variables.

Monthly blood samples were collected over one year in 10 elite cyclists (25.6 ± 3.4 yrs, 181 ± 4 cm, 71.3 ± 4.9 kg, 6.7 ± 0.8 W.kg^-1^ 5-min maximal power output) to calculate individual ABP profiles and monitor hematological variables. Total hemoglobin mass (Hbmass) and PV were additionally measured by carbon monoxide rebreathing. Acute and chronic training loads – respectively 5 and 42 days before sampling – were calculated considering duration and intensity (training stress score, TSS™).

[Hb] averaged 14.2 ± 0.0 (mean ± SD) g.dL^-1^ (range: 13.3 to 15.5 g·dl^-1^) over the study with significant changes over time (*P* = 0.004). Hbmass was 1’030 ± 87 g (range: 842 to 1116 g) with no significant variations over time (*P* = 0.118), whereas PV was 4309 ± 350 mL (range: 3688 to 4751 mL) with a time-effect observed over the study time (*P* = 0.014). Higher acute – but not chronic – training loads were associated with significantly decreased [Hb] (*P* <0.001). Although individual hematological variations were observed, all ABP variables remained within the individually calculated limits.

Our results support that acute training load variations significantly affect [Hb], likely due to short-term PV fluctuations, underlining the importance of considering training load when interpreting individual ABP variations for anti-doping purposes.

## 1 Introduction

To prevent blood doping in elite cycling, in 2008 the Union Cycliste Internationale (UCI) spearheaded the introduction of the Athlete Biological Passport (ABP) (Zorzoli et al., 2010). The World Anti-Doping Agency (WADA) then progressively implemented the ABP more widely and currently more than 30’000 blood samples are collected yearly to longitudinally track various blood markers of athletes (Faiss et al., 2020;Saugy et al., 2020). Starting with average population levels as initial reference, biomarkers in successive samples from a given athlete allow an individually expected range to be predicted within which the series of marker values should fall assuming physiological conditions (WADA, 2019a). This range is calculated with an adaptive Bayesian statistical model using levels of probability (i.e. specificity) chosen to estimate the limits of normal physiological variation (Sottas et al., 2011). The premise is that repeated sampling allows for a progressive narrowing of the range of values considered as physiological for a given individual. The adaptive model uses hemoglobin concentration ([Hb]) and a stimulation index, the OFF-score (combining reticulocyte percentage (Ret%) and [Hb]), to generate an Atypical Passport Finding (ATPF) if a marker falls outside the expected range with a 99% specificity (i.e. 1:100 chance or less that this result is due to normal physiological variation) (WADA, 2019a).

The individualized ranges for the ABP variables need to be sufficiently large and robustly defined to avoid an ATPF caused by fluctuations related to factors independent of blood doping (Sottas et al., 2011). Several potentially confounding factors have been investigated for their putative influence on ABP variables, beyond natural physiological variations. First, blood sample quality is paramount as there is a potential influence of pre-analytical procedures (e.g. position during sampling, sample transport) on the analytical results (Ahlgrim et al., 2010;Robinson et al., 2011;Ashenden et al., 2014;Astolfi et al., 2020). There are strict WADA guidelines for blood collection, transportation and storage to prevent variations due to such confounders (WADA, 2019b;a;c). As a further improvement an easy-to-use blood sample quality index (blood stability score (BSS)) is currently also implemented (Robinson et al., 2016).

Some markers of the ABP may be influenced by plasma volume (PV) variations altering their concentration in whole blood. Exposure to extreme environments (e.g. hot or hypoxic) can alter PV and [Hb] (Sawka et al., 2000;Stanley et al., 2015;Lobigs et al., 2018a;Young et al., 2019;Coffman et al., 2020). Blood loss, injury or illness may have an effect on total hemoglobin mass (Hbmass) and [Hb]. Since such variations alter the ABP profiles (Gough et al., 2013) these confounders should be mentioned on the blood control forms (WADA, 2016) to allow for an informed evaluation of the ABP. In response to a call for inclusion of “all other relevant information also comprising training and competition results” (Vernec, 2014), monitoring athletic performance (and hence training content) has been proposed (Faiss et al., 2019), to further strengthen the ABP and its interpretation. Hematological biomarkers vary during a competitive season in athletes among disciplines (Banfi et al., 2006;Banfi et al., 2011;Diaz et al., 2011;Andelkovic et al., 2015). This could be due to plasma volume variations induced by effort in competition, or strength and endurance training periods (Collins et al., 1986;Imelik et al., 1992;Sawka et al., 2000). Little is known about any direct influence of training load variation on hematological variables (Guglielmini et al., 1989;Varamenti et al., 2018). There are no studies investigating the influence of training load over a prolonged period on the ABP variables in elite cyclists.

The purpose of this study was therefore to monitor training load in elite cyclists over one year and analyze if and how individual ABP profiles constructed from monthly blood samples vary with training load. To further address the within-subject variance of [Hb] as a primary marker of the ABP (Lobigs et al., 2016;Garvican-Lewis et al., 2020), we also measured directly PV and Hbmass to assess whether changes due to environmental conditions (seasonal effect) or prolonged periods of high vs. low training loads alter PV and Hbmass to an extent affecting ABP profiles. We hypothesized that acute (5 days) and chronic (42 days) training before an ABP sample would notably alter the profile readings without exceeding the individual ranges of the ABP adaptive model.

## 2 Materials and methods

### 2.1 Study participants

Ten elite cyclists (25.6 ± 3.4 yrs, 181 ± 4 cm, 71.3 ± 4.9 kg, 6.7 ± 0.8 W.kg^-1^ 5-min maximal power output) volunteered to participate in the study. All were members of the Swiss national cycling team or an elite cycling team registered at Swiss Cycling and competing in road, track and mountain-bike cycling events at an international level (e.g. UCI World & Europe Tour races or UCI World Cups). They lived < 800 m and were healthy at the start of the study. Initially, 12 subjects were recruited. One subject withdrew due to personal reasons. Another subject was excluded because of a medical condition during the study affecting his hematological variables, precluding the training load from being considered as the major factor of any variation in the hematological variables. All participants provided a fully informed written consent to participate after the procedures and risks were explained. The study protocol was approved by the regional research ethics committee (CER-VD, Lausanne, Switzerland, #2018-01019) and conducted in respect of the Declaration of Helsinki.

### 2.2 Blood sampling and analysis

Venous blood samples were collected monthly from every participant by the same experienced phlebotomist. Due to competition schedules and training camps, the samples were separated by 32 ± 12 days. WADA blood collection guidelines were strictly followed with no physical exercise allowed in the 120 min preceding sampling and blood collection done after 10 min in a seated position (WADA, 2016;2019a). Venipuncture was realized with a 21G short manifold butterfly needle inserted into an antecubital vein (Sarstedt Safety-Multifly®, Sarstedt AG, Nümbrecht, Germany), and blood was collected in EDTA-coated tubes (Sarstedt S-Monovette EDTA-K 2.7 mL, Sarstedt AG, Nümbrecht, Germany). Samples were stored at 4° C for 30 min to 12 h after collection before analysis, depending on instrument and technician availability. Samples were homogenized at room temperature (21° C) on a roller system for 15-45 min before analysis with a fully automated flow cytometer (Sysmex XN1000, Sysmex Europe GmbH, Norderstedt, Germany). Internal quality controls provided by the manufacturer (Sysmex E-Checks, levels 1, 2, and 3) were run three times before each batch of samples. The analysis was repeated to produce two successive analyses with differences equal or less than 0.1 g.dL^-1^ for [Hb], and 0.15% or 0.25% for Ret% (depending whether Ret% was inferior or superior to 1%) conforming to the applicable WADA guidelines (WADA, 2019a). The first valid test result was then recorded. The stimulation index OFF-score was calculated as ([Hb] x 10) - 60 x √Ret% and the Abnormal Blood Profile Score (ABPS) was calculated combining Ret%, [Hb], hematocrit (HCT), red blood cell number (RBC#), mean red cell volume (MCV), mean red cell Hb (MCH) and mean cell Hb concentration (MCHC) (WADA, 2019b).

### 2.3 Individual ABP profiles

For each participant an individual longitudinal ABP profile was constructed with the values obtained from the collected blood samples using the official training version of the ABP-module in WADA’s Anti-Doping Administration and Management System (ADAMS). The system calculates ABPS and OFF-score for each sample and then generates individual ABP profiles with [Hb] and OFF-score as primary markers and Ret% and the ABPS as secondary ones. Population-based upper and lower limits are used for the first blood sample after which an adaptive model generates individually varying limits for each subsequent blood sample considering previous individual analytical results. An Atypical Passport Finding (ATPF) is generated when a) a [Hb] and/or OFF-score value of the last entered sample falls outside the lower and upper intra-individual limits or b) when the last 2 to 5 [Hb] and/or OFF-score values deviate from the expected range (a so-called “sequence ATPF”). For the first case, the applied specificity is 99% (i.e. 1:100 chance or less that the deviation is due to normal physiological variation). For the latter, the applied specificity is 99.9% (i.e. 1:1000 chance or less that the sequence deviation is due to normal physiological variation). An ATPF results in a notification to the Athlete Passport Management Unit (APMU) handling the administration of the individual passport on behalf of a passport custodian. The APMU may request expert opinions and declare an adverse passport finding (APF) after 3 independent experts with all available information unanimously deemed the profile likely to result from doping. An APMU may also request an expert opinion in the absence of an ATPF when abnormal variations (e.g. compatible with artificial hemodilution) is observed with ABP profiles remaining within individual limits.

To complement the analysis of variations in ABP variables, for [Hb] and the OFF-score, we also calculated the shortest absolute distance to the closest individual limit (i.e. from the upper or the lower limit).

### 2.4 Total hemoglobin mass and plasma volume

Hbmass was determined monthly over the last eight months of the study with a fully automated blood volume analyzer (OpCo: Detalo Instruments, Birkerod, Denmark) based on a carbon monoxide (CO) rebreathing technique, as described elsewhere (Siebenmann et al., 2017). Briefly, participants were comfortably installed in supine position with a nose clip and a mouthpiece connected to a closed rebreathing circuit. They then breathed 100% oxygen (O_2_) for 4 min to flush the airways of nitrogen. Subsequently, a bolus of 1.5 mL/kg of 99.997% chemically pure CO (Carbagas, Liebefeld, Switzerland) was introduced into the circuit after which the participants rebreathed the O_2_-CO mixture for 9 minutes. Venous blood was then drawn from an antecubital vein and immediately analyzed in triplicate for carboxyhemoglobin content (HbCO%) with a calibrated gasometer (ABL80-Co-Ox, Radiometer, Copenhagen, Denmark). Initial duplicate measurements in our laboratory yielded a typical error (TE) of 1.8%, in line with previously reported values (Siebenmann et al., 2017;Rønnestad et al., 2020). The CO remaining in the system was measured with a CO meter (Monoxor Plus, Bacharach, New Kensington, USA) and subtracted from the initial amount introduced to define the exact CO bolus received with a 0.1 mL typical error. Hbmass was calculated from the difference in HbCO% before and after CO-rebreathing. Total red blood cell volume (RBCV), plasma volume (PV) and blood volume (BV) were derived from [Hb], hematocrit (HCT) and Hbmass.

### 2.5 Training load quantification

Participants were instructed to follow their habitual training and competition schedules as planned with their personal trainer and to report all their training and competition activities in a commercially available online training monitoring interface (Training Peaks™ (TP), PeaksWare, Lafayette, CO, USA). Since all participants were already using TP to monitor their training, we could collect training data for the 42 days prior to the first blood sampling in addition to the twelve months of the monitoring of their hematological variables. All used a crank-based power meter (SRM, Schoberer Rad Messtechnik, Juelich, Germany) for their cycling-based training sessions allowing their training load to be accurately quantified as a function of the duration and intensity of each training session. They were instructed to proceed to regular static calibration and zero-offset calibrations of their power meter according to the manufacturers’ recommendations.

The Training Stress Score (TSS™, arbitrary units) was automatically calculated for each training session in TP using the following formula: TSS (a.u.)= [(*t* x *NP* x *IF*) / (*FTP* x 3600)] x 100 where *t* is the duration in seconds, FTP represents the functional threshold power calculated as 95% of the average power from a recent 20-minute steady-state all-out time trial or maximal effort, *NP* is the normalized power, representing a calculation of the power that could have been maintained for the same physiological “cost” if the power had been perfectly constant, and *IF* is the intensity factor indicating the relative intensity of the session calculated as the ratio of NP to FTP (Coggan, 2019).

Individual FTP values were determined at the beginning of the study based on the results of a maximal 20-min field or laboratory test realized under supervision of their personal trainer. The initially calculated FTP was not modified during the study to allow for an adequate comparison of training loads and their variation throughout the study.

Acute training load (ATL) was calculated as the load during the 5 days preceding blood sampling both as a score cumulating and averaging the TSS over 5 days. Chronic training load (CTL) was defined as the load for the 42 days (6 weeks) preceding blood sampling and calculated again as a cumulated and averaged TSS over the period.

Training loads for High vs. Low training load periods for each cyclist were obtained by identifying the periods with the highest and lowest 12-week cumulative TSS. Seasonal training variation (Winter vs. Summer) was quantified calculating the cumulated TSS during three winter months (December, January and February) and three summer months (June, July, August), respectively.

### 2.6 Time-trial performance assessment

Aerobic endurance performance was assessed after five months of data collection with a 250kJ time-trial (Stevens et al., 2015). Using their own bicycle fixed on a stationary trainer equipped with a calibrated power meter (Tacx, Neo Smart, T2800, Wassenaar, The Netherlands), cyclists were requested to reach 250 kJ as fast as possible, while average power output and completion time were recorded. Briefly, participants warmed up for 10 min at a self-selected pace, and then rested seated on their bicycle for 5 min before the time-trial started. The trainer was connected to a computer running dedicated software (Tacx Software, Tacx, Wassenaar, The Netherlands) simulating a flat outdoor time-trial. Only the incremental mechanical work (kJ) completed was displayed on the screen.

### 2.7 Statistical analyses

Values are reported as means and standard deviations. Using data from 12 monthly blood samples for each participant (cluster variables, random factor), repeated measures analyses were conducted for each of the four primary variables of the ABP (i.e. [Hb], Ret%, OFF-score and ABPS) with a mixed model to determine whether changes in the dependent variables (ABP variables) differed over time (fixed factor). This technique was preferred to a repeated measures ANOVA because it also allows to handle dynamic predictors. The effect on the ABP variables, Hbmass, and plasma volume of acute or chronic training load, seasonal variations in training load as well as periods with High vs. Low training load were assessed with mixed models using training load as time-dependent covariates. Visual inspection of residual plots allowed excluding any obvious deviations from homoscedasticity or normality. Polynomial contrasts were used for time in mixed models, employing the Bonferroni method. The Pearson correlation coefficient was calculated for the relationship between time-trial performance (s) and Hbmass. The level of significance was set at *P* < 0.05. All statistical analyses were conducted with an open source dedicated statistical software (Jamovi, Jamovi Project Software, retrieved from https://www.jamovi.org).

## 3 Results

### 3.1 Hematological variations over 12 months

[Hb] averaged 14.2 ± 0.1 g·dl^-1^ (range: 13.3 to 15.5 g.dL^-1^) over the study, while the ABPS averaged -1.32 ± 0.41 a.u. (range: -1.68 to -0.40 a.u.), both with significant variations over the 12 months ([Hb] (F (11, 99) = 2.76, *P* = 0.004) and ABPS (F (11, 99) = 3.54, *P* < .001). Ret% averaged 1.2 ± 0.31 % (range: 0.76 to 1.81 %) and the OFF-score 76.6 ± 10.0 a.u. (range: 64.0 to 91.7 a.u.) with no significant variation over the study time (Ret% (F (11, 99) = 1.85, *P* = 0.056) and OFF-score (F (11, 99) = 1.77, *P* = 0.069). Average hematological values over the 12 months for each cyclist are presented in Table 1. Hbmass averaged 1030 ± 87 g (range: 842 to 1116 g) with no significant variation over the study time (F (7, 52) = 1.75, *P* = 0.118). Conversely, a significant time effect was observed for PV (F (7, 52) = 2.83, *P* = 0.014) which averaged 4309 ± 350 mL (range 3688 to 4751 mL).

**Table 1.** Average individual hematological variables and training load over 12 months. Values reported as means ± SD. [Hb]: hemoglobin concentration; OFF-score; Ret%: reticulocytes percentage; ABPS: abnormal blood profile score; Hbmass: total hemoglobin mass; PV: plasma volume; along with acute training load (5 days, ATL), chronic training load (42 days, CTL) before blood sampling and total training stress score (TSS) over one year; a.u.: arbitrary units. *: Hbmass and PV were monitored during the last eight months of the study.

|  | [Hb]<br>g/dl | OFF-score<br>a.u. | Ret<br>% | ABPS<br>a.u. | Hbmass*<br>g | PV*<br>mL | ATL<br>a.u. | CTL<br>a.u. | Total TSS<br>a.u. |
| --- | --- | --- | --- | --- | --- | --- | --- | --- | --- |
| Cyclist 1 | 14.4 ± 0.4 | 91.7 ± 4.7 | 0.76 ± 0.1 | -1.47 ± 0.2 | 1021 ± 45 | 4222 ± 225 | 63.4 ± 55 | 53 ± 27 | 18871 |
| Cyclist 2 | 13.5 ± 0.8 | 72.9 ± 8.3 | 1.07 ± 0.2 | -1.68 ± 0.3 | 1021 ± 51 | 4703 ± 491 | 48.8 ± 50 | 73 ± 22 | 26814 |
| Cyclist 3 | 13.8 ± 0.5 | 65.3 ± 6.5 | 1.49 ± 0.2 | -1.59 ± 0.3 | 1045 ± 28 | 4751 ± 185 | 51.2 ± 55 | 61 ± 17 | 22466 |
| Cyclist 4 | 15.5 ± 0.6 | 86.9 ± 5.8 | 1.31 ± 0.2 | -0.40 ± 0.5 | 1157 ± 24 | 3998 ± 641 | 22.1 ± 38 | 36 ± 28 | 16751 |
| Cyclist 5 | 14.8 ± 0.4 | 85.2 ± 4.4 | 1.08 ± 0.2 | -0.91 ± 0.5 | 1014 ± 33 | 3944 ± 374 | 105 ± 63 | 79 ± 36 | 30957 |
| Cyclist 6 | 13.3 ± 0.6 | 64.4 ± 11.0 | 1.32 ± 0.4 | -1.59 ± 0.2 | 950 ± 35 | 4318 ± 377 | 117 ± 54 | 125 ± 25 | 48050 |
| Cyclist 7 | 13.4 ± 0.4 | 80.3 ± 5.7 | 0.8 ± 0.2 | -1.58 ± 0.1 | 842 ± 17 | 3688 ± 208 | 47.5 ± 36 | 61 ± 19 | 19439 |
| Cyclist 8 | 14.3 ± 0.6 | 74.1 ± 7.0 | 1.32 ± 0.2 | -1.55 ± 0.3 | 1116 ± 21 | 4637 ± 338 | 104 ± 47 | 95 ± 11 | 38090 |
| Cyclist 9 | 14.2 ± 0.5 | 80.9 ± 5.5 | 1.05 ± 0.1 | -1.52 ± 0.4 | 1079 ± 18 | 4417 ± 286 | 77.5 ± 65 | 103 ± 14 | 33223 |
| Cyclist 10 | 14.4 ± 0.6 | 64.0 ± 5.7 | 1.81 ± 0.3 | -1.04 ± 0.5 | 1056 ± 33 | 4421 ± 309 | 71.5 ± 57 | 104 ± 32 | 36620 |
| Mean | 14.2 ± 0.01 | 76.6 ± 10.0 | 1.2 ± 0.31 | -1.32 ± 0.41 | 1030 ± 87 | 4309 ± 350 | 70.8 ± 30 | 79 ± 27 | 29128 ± 10091 |

### 3.2 Within-subject variations

The measured variables in the ABP profiles remained within the individualized limits; and no ATPFs were outlined. The distance to the individual limits for [Hb] and OFF-score values yielded an average mean lowest distance to the limits of 1.0 ± 0.4 g·dL^-1,^ and 18.5 ± 3.9 (a.u.), respectively. Over the 120 measurements (10 subjects x 12 blood samples), 10 [Hb] values (8.3%) fell within a distance to the (upper or lower) individual limit < 0.5 g·dl^-1^ and 4 within a < 0.1 g·dl^-1^ (3.3%) distance. Four individually calculated OFF-score values were closer than 5 points (a.u.) to an individual limit (3.3%). Three illustrative examples of individual ABP profiles are presented in Figures 2, 3 and 4; and the hematological profiles of the remaining cyclists are available as Supplementary files.

**Figure 1.**
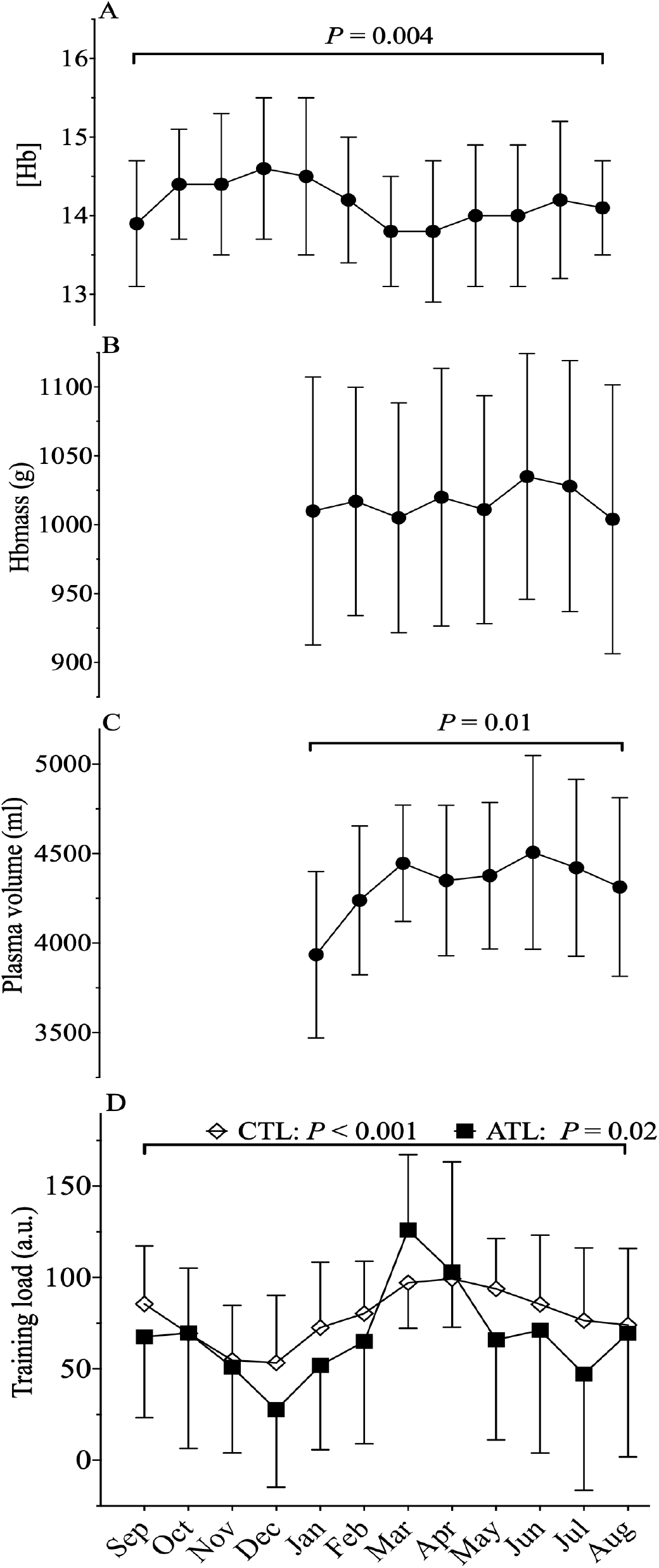
Average 12 months representation of **(A)** [Hb]: hemoglobin concentration; **(B)** Hbmass: total hemoglobin mass; **(C)** PV: plasma volume; along with **(D)** training load (a.u.) (average daily TSS) with acute training load (5 days, ATL) and chronic training load (42 days, CTL) before sampling; a.u.: arbitrary units. *P* – values for the statistical difference over the 12 months.

**Figure 2.**
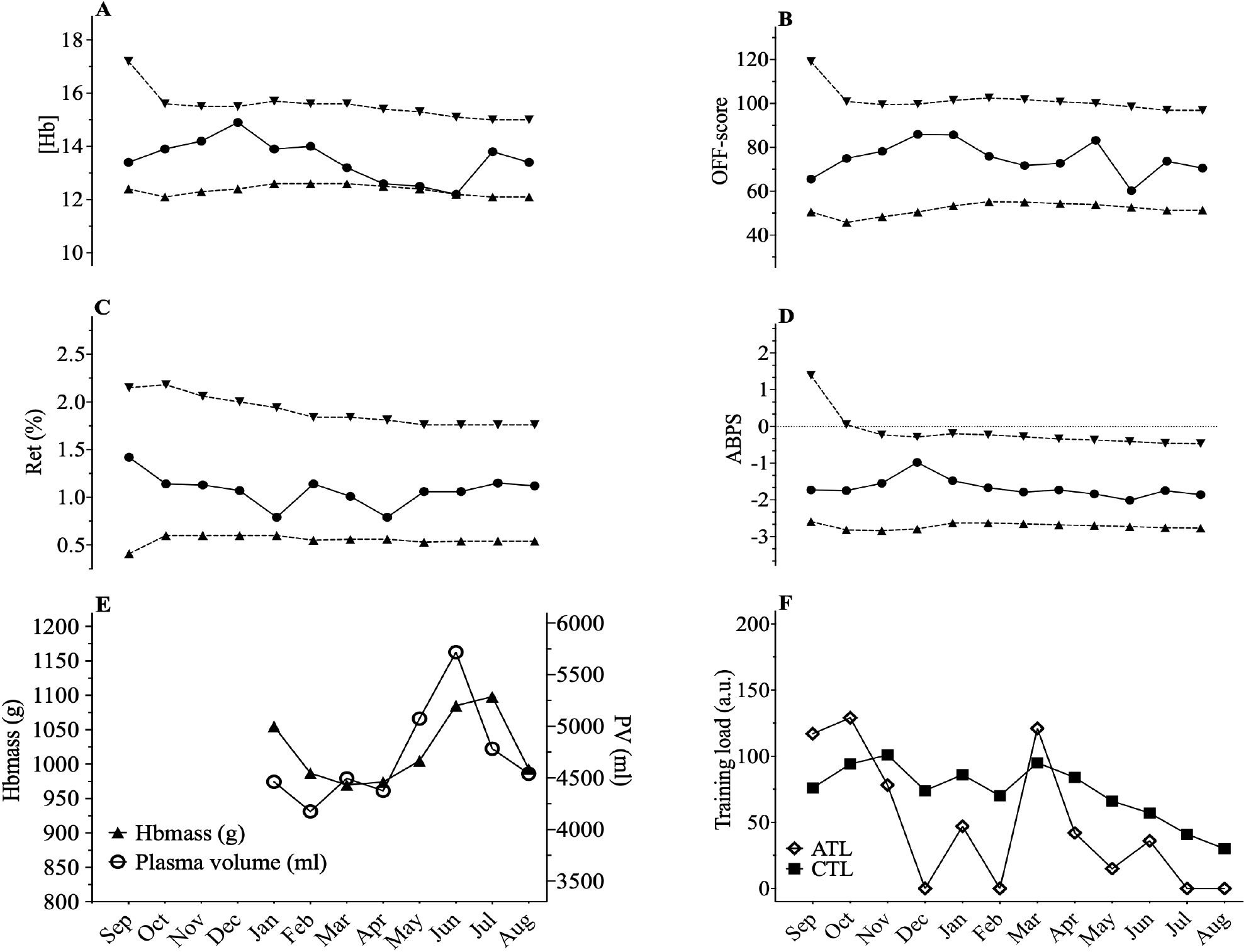
ABP profile for cyclist 2 showing a prolonged state near the individual limits calculated for [Hb]. [Hb] decreased from December to June by 17.6 % however, an increase in Hbmass of 3% between January and June was observed with an increase of 28% of PV (4461-5719 mL) despite a decrease in chronic training load of -65% from January to August.

**Figure 3.**
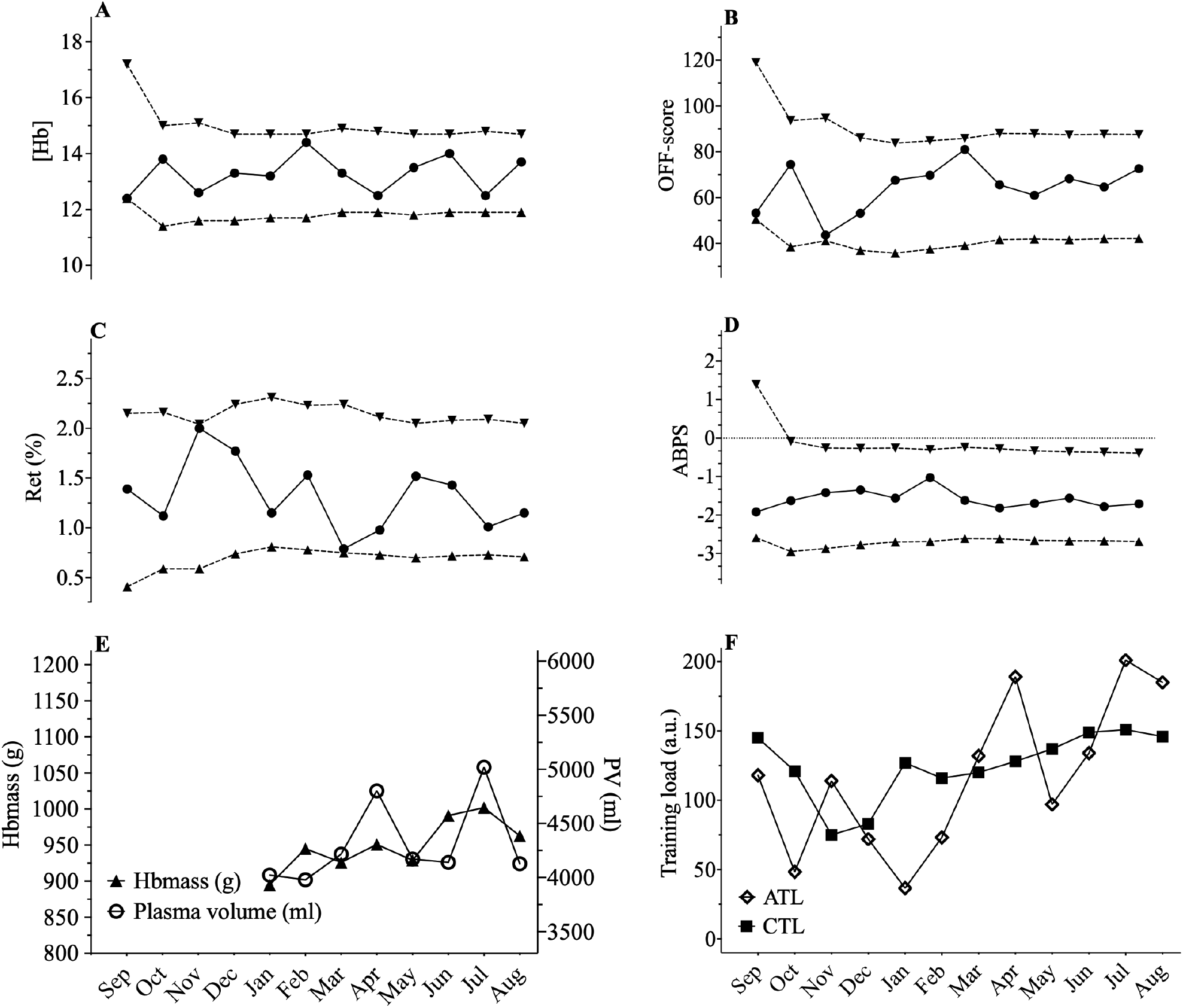
ABP profile for cyclist 6 showing high variability in hematological parameters, especially for [Hb]. Mean [Hb] was 13.5 ± 0.77 g.dL^-1^ for this cyclist and represents the highest SD among all the cyclists with values ranging from 12.2 to 14.4 g.dL^-1^ t (variation of 16%). Limits values are also encountered for OFF-score and Ret%. However, Hbmass varied only by 2% between February until August (945-963 g) and only 3% variations in PV for the same period (3979-4129 mL). Despite high variations in the ABP biomarkers, chronic training load did not significantly vary for this cyclist from January to August (+15%).

**Figure 4.**
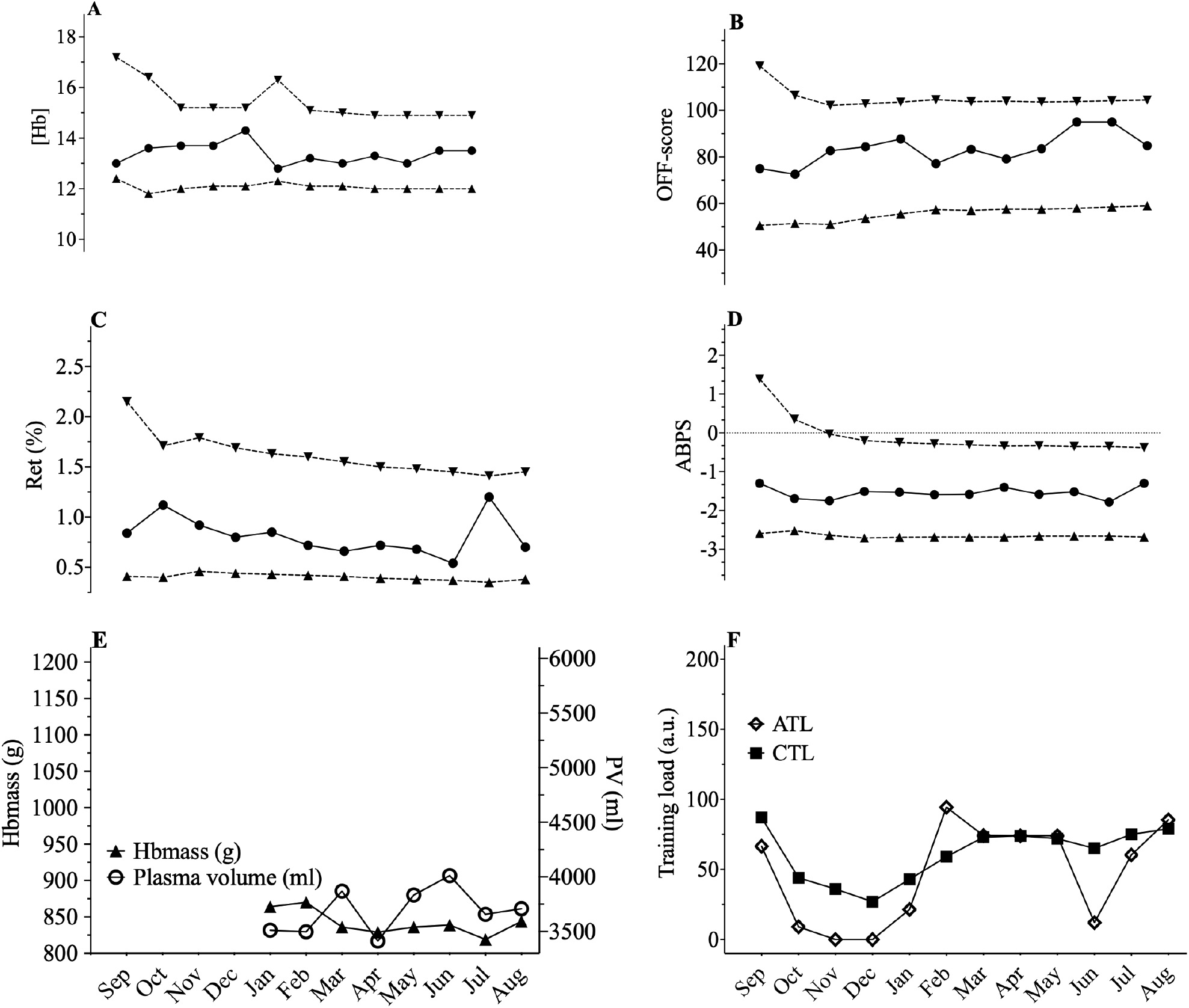
ABP profile for cyclist 7 showing few variations. [Hb] varied from 12.8 to 14.4 (10%) with a 6% Hbmass variation (819 to 870 g) despite a 18% change in PV (3413 mL to 4012 mL) and a chronic training load increase of 83% from January to August.

Figures 2-3-4. Representations of the Athlete Biological Passport (ABP) hematological profile for cyclist 2 (Figure 2), cyclist 6 (Figure 3), and cyclist 7 (Figure 4) with (A) [Hb]: hemoglobin concentration; (B) OFF-score, (C) Ret%: reticulocytes percentage; (D) ABPS: abnormal blood profile score; over the 12 months of the study design. Solid lines represent the athlete’s values, dotted lines represent the upper limits and lower limits calculated by an adaptive Bayesian model (see methods section for details). (E) Acute training load (ATL) and chronic training load (CTL) represent the load respectively 5 and 42 days before sampling (F). Triangles figure total hemoglobin mass (g) and circles represent plasma volume (mL) over the last eight months of the study.

### 3.3 Training load analysis

The average cumulated TSS over 12 months amounted to 29128 ± 10091 a.u.. For the High load period average cumulated TSS amounted to 10389 ± 2933 a.u. vs. 3440 ± 2544 a.u. during the Low load period. Cumulated TSS during the Low load period represented on average 31 ± 18% of the High Load TSS. The average cumulated Winter TSS was 7396 ± 2817 a.u., representing on average to 89 ± 50% of the Summer TSS (7609 ± 3658 a.u.) *P* = 0.75) (Table 2). Cumulated TSS was 353 ± 292 a.u., and 3310 ± 1466 a.u over the 5 and 42 days preceding blood sampling, respectively. Average mean daily TSS over the 5 days preceding blood sampling (ATL, 71 ± 30 a.u.) and average mean of daily TSS over the 42 days preceding blood sampling (CTL, 79 ± 27 a.u.) varied significantly with time (*P* = 0.02 for ATL and *P* < 0.001 for CTL) (Figure 1 and Table 1).

**Table 2.** Average hematological variables according to season and to training load. Data are presented as means ± SD. [Hb]: hemoglobin concentration, OFF-score, Ret%: reticulocytes percentage; ABPS: abnormal blood profile score; Hbmass: total hemoglobin mass; PV: plasma volume; a.u.: arbitrary units. Summer vs. winter were calculated as the cumulated TSS over three months, high vs. low training loads are calculated as the maximal and minimal cumulated TSS per months over three successive months. * Hbmass and PV were monitored during the last eight months of the study. ^#^ significantly different from Low load (TSS) P < .001.

|  | Summer | Winter | Change (%) | High load | Low load | Change (%) |
| --- | --- | --- | --- | --- | --- | --- |
| [Hb] (g/dl) | $14.1 \pm 0.8$ | $14.2 \pm 0.7$ | 0.7 | $14.0 \pm 0.6$ | $14.4 \pm 1.0$ | 2.8 |
| Off-score (a.u.) | $76.8 \pm 10.0$ | $78.8 \pm 11.1$ | 2.6 | $77.1 \pm 9.3$ | $77.6 \pm 11.9$ | 0.6 |
| Ret% | $1.18 \pm 0.3$ | $1.15 \pm 0.3$ | -2.5 | $1.1 \pm 0.3$ | $1.3 \pm 0.3$ | 18.2 |
| ABPS (a.u.) | $-1.42 \pm 0.5$ | $-1.25 \pm 0.4$ | 12.0 | $-1.44 \pm 0.4$ | $-1.23 \pm 0.6$ | 14.5 |
| Hbmass (g)* | $1032 \pm 84$ | $1023 \pm 86$ | 0.9 | $1048 \pm 99$ | $1079 \pm 41$ | 3.0 |
| PV (mL)* | $4442 \pm 403$ | $4148 \pm 393$ | -6.6 | $4399 \pm 354$ | $4342 \pm 502$ | -1.3 |
| TSS (a.u.) | $7609 \pm 3658$ | $7396 \pm 2817$ | -2.8 | $10389 \pm 2933^{\#}$ | $3440 \pm 2544$ | -67 |

#### 3.3.1 Seasonal and training load period influence on the ABP

Training load period (High load vs. Low load) did not significantly affect ABP variables (i.e., [Hb] (*P* = 0.50), OFF-score (*P* = 0.49), Ret% (*P* = 0.16), ABPS (*P* = 0.29)) or Hbmass (*P =* 0.36) and PV (*P* = 0.86). There was no significant effect of season on the ABP variables (i.e., [Hb] (*P* = 0.87), OFF-score (*P* = 0.96), Ret% (*P* = 0.87), ABPS (*P* = 0.20) and on Hbmass (*P* = 0.45) and PV (*P* = 0.06). Hematological variations with respect to High vs. Low training load periods and Winter vs. Summer are summarized in Table 2.

#### 3.3.2 Influence of acute and chronic training load on the ABP variables

There was a significant effect of ATL on [Hb] (F (1,102) = 12,8, *P* < .001), b = - 0.0036, and PV (F (1,55) = 7.76, *P* = 0.007, b= 2.2, but not ABPS ((F (1,102) = 3.35, *P* = 0.07), OFF-score ((F (1, 102) = 3.77, *P* = 0.055), Ret % ((F (1, 102) = 1.18, *P* = 0.28 or Hbmass ((1, 52) = 2.05, *P* = 0.16). No significant effect of CTL was observed neither on the ABP variables nor Hbmass and PV ([Hb] (F (1,106) = 1.86, *P* = 0.176), Ret% (F (1,106) = 1.62, *P* = 0.2), OFF-score (F (1, 106) = 0.016, *P* = 0.89), ABPS (F (1, 106) = 2.34, *P* = 0.129), Hbmass (F (1,55)= 1.72, *P* = 0.195), PV (F (1, 57)= 0.85, *P* = 0.36). For each cyclist, ATL, CTL and total training load over the 12 months are reported in Table 1.

Cumulated TSS over the 42 days preceding blood sampling had no effect on [Hb] (F (18,82) = 1.19, *P* = 0.35), OFF-score (F (18, 82) = 1.17, *P =* 0.36, Ret% (F (17.3,82) = 1.87, *P* = 0.071), ABPS (F (16.7,82) = 2.02, *P* = 0.053), Hbmass (F (2,54) = 1.29, *P* = 0.53), PV (F (2,54) = 1.99, *P* = 0.39), Cumulated TSS over the 5 days preceding blood sampling had a significant effect on [Hb] (F (1,101) = 12.91, *P* < .001), b = -7.35e-4, OFF-score (F (1, 100) = 4.32, *P =* 0.04), b = - 0.00545 and PV (F (1,56) = 7.76, *P* = 0.007), b = 0.44 There was no significant effect on Ret% (F (1,100) = 0.79, *P* = 0.38), ABPS (F (1,101) = 3.20, *P* = 0.08), PV (F (2,56) = 18.5, *P* = 0.053) and Hbmass (F (1,52) = 2.05, *P* = 0.16)

#### 3.3.3 Time-trial performance

Average time-trial duration was 668 ± 40 s for an average power of 393 ± 42 W. Time trial performance was correlated with absolute Hbmass (r = 0.75, *P* = 0.02) but not when Hbmass was expressed relative to body mass (r = -0.55, *P* = 0.12).

## 4 Discussion

By collecting and analyzing monthly blood samples together with training stress score in a cohort of elite cyclists over a one-year period, we could test the hypothesis that variations in training load over time lead to relevant changes in ABP parameters. The main finding of this study was that acute changes in training load (5 days) prior to blood sampling influenced ABP parameters (e.g., [Hb]), but not chronic changes in training load (42 days). We observed significant variations in PV (but not Hbmass) over time. Despite the highlighted variations ABP variables remained within the individual limits at all times.

Endurance athletes reportedly have greater PV in comparison with team sports athletes, power endurance athletes, and disabled or untrained subjects (Fellmann, 1992). Endurance athletes also have fluctuations in PV, potentially inducing variations in biological markers that are concentration sensitive such as [Hb] (Lobigs et al., 2016). In agreement we found [Hb] and PV to significantly vary over a one-year period, while Hbmass did not. Monitoring PV would hence allow to interpret [Hb] alterations adequately. Variations in environmental temperature may also affect blood variables (Sawka et al., 1987). We found for instance that a training period in warm summer months was associated with a significant 4.7 % increase in PV (when compared to winter) even though ABP variables or Hbmass were not influenced. Interestingly, we found that, despite PV discrepancies (Summer vs Winter), these were not due to significant differences in the training load between those two periods (Table 2).

Our study allowed to contrast the influence of acute (5 days, ATL) variations of training load with the load considered over a longer time (42 days, CTL) before each blood sample. We found that higher ATL was accompanied by lower [Hb] and increased PV. This strongly suggests a hemodilution associated with short-term ATL fluctuations. An acute hemodilution was recently observed in professional floorball players immediately after a game (>3% decrease in [Hb]) while values returned to baseline after 2 h (Wedin et al., 2020). In an anti-doping context, the pre-analytical bias possibly due to the acute effect of one single strenuous effort is avoided with the compulsory 2-h waiting time after the exercise before blood sampling is allowed (WADA, 2019b). The latter rule does however not apply to repeated exercises (i.e. training load) on the days before a blood sample. Our results therefore suggest that monitoring or at least reporting the training load for several days before blood sampling would reasonably allow a better interpretation of ABP variables in an anti-doping setting.

Intuitively, phases with possibly lower training loads (e.g., holidays or off-season periods) are expected to have an influence on blood variables. In our cohort of elite cyclist, we identified prolonged three-months periods with significantly higher training loads, but these did not have a significant effect on the variation of ABP profiles (Table 2). Conversely, [Hb] was shown to decrease in a workload dependent manner while red blood cell count remained constant in 19 elite competitive soccer players over half a competitive season of three months with a controlled training program (Andelkovic et al., 2015). This underlines the prime relevance of within-subject variation in the ABP that needs to be considered on an individual basis, despite the scientific relevance of cohort results.

The ABP was hence designed to allow “switching the focus from comparison with a population to the determination of individual values” (Sottas et al., 2010). Bayesian networks were used for the ABP, because they allow to represent the causal relationship between blood doping and its effect on hematological biomarkers (Koski et al., 2009;Kruschke, 2011). For instance, if blood doping (e.g., recombinant human erythropoietin (rhEPO) use) leads to an increased [Hb], rhEPO is the cause and a higher [Hb] the effect. Monitoring hematological biomarkers in a longitudinal profile is thus challenging because it goes against the causal direction. The way the ABP was designed allows however to analyse the probability that hematological variations may be due to doping rather than natural fluctuation based on existing data showing reference ranges and within-subject variability of either doped or non-doped populations (Malcovati et al., 2003). More precisely, the model relies i) on a dichotomic variable with two states: doped or non-doped, and ii) a continuous variable represented by hematological biomarkers. Bayes’ theorem then expresses the probability of being in a doped state as a function of the measured biomarker. Individual limits of each biomarker of the ABP are set with a high specificity (e.g., 99%). This means that there is less than 1:100 chance that a value outside of the limits is due to a normal physiological condition. The advantage of Bayesian Networks is that they allow to include heterogeneous and confounding factors (e.g., age, sex, ethnic origin, type of sport, altitude exposure) (Sottas et al., 2010). Already when it was launched, the potential of the ABP in integrating new potential confounding factors (i.e. training load) to the Bayesian adaptive model was acknowledged (Sottas et al., 2011). Now the inclusion of performance models has also been proposed (Faiss et al., 2019).

Besides, biomarkers of PV variations were also proposed to improve the ABP adaptive model by removing the confounding effect of PV variations (Lobigs et al., 2018b;Garvican-Lewis et al., 2020). More importantly their results showed that including PV correction for the calculation of primary ABP markers (i.e. [Hb] and OFF-Score) increases sensitivity (i.e. the probability to find a true positive) without any loss of specificity (i.e. the true negative rate). By applying the PV correction, 66% of [Hb] within-subject variance could be explained (Lobigs et al., 2018b). In our study, to complement the ABP approach, we addressed within-subject variance, and the influence of PV variations by looking at the lowest distance to the individual limits calculated by the Bayesian model for each successive sample. For example, we observed 3 successive [Hb] values within 0.1 g·dL^-^ to the individual limit with a concomitant increase in PV of 1344 mL in one cyclist (Figure 2). However, when considering all 10 elite cyclists (120 ABP points over one year) and despite noticeable differences in PV or [Hb], no ATPFs were observed, with the lowest distance to the individualized upper and lower limits for [Hb] falling only 10 times < 0.5 g·dL^-1^. The inspection of individual ABPs however revealed various patterns, with variations potentially related or unrelated to training. These variations were not uniform and individual interpretation of within-subject variance in the context of the APB is paramount: a decrease in [Hb] may result from a large increase in PV (over 1200 mL), likely related to increased training load (see example in Figure 2). Alternatively, changes in PV or Hbmass may not necessarily result in concomitant fluctuations in [Hb] despite a change in training load (see example in Figure 3). Finally, a rather stable ABP profile may appear despite high variations in PV (+ 28%) and Hbmass (+ 6%) (see Figure 4). The latter example would additionally question the usefulness of including training content in the interpretation of a profile with no noticeable [Hb] variation notwithstanding significant changes in training load. To summarize, despite statistically significant relations obtained on aggregate data, there was no systematic association between PV, Hbmass, and individually interpreted ABP variables. Overall our results suggest that the current individual limits of the ABP seem sufficiently robust to prevent a falsely negative interpretation of an ABP profile even though training load variations are present.

Bearing this in mind, blood doping remains attractive to augment Hbmass and improve convective oxygen transport capacity (Warburton et al., 2000) even with low-volume transfusions that can have a significant performance enhancing effect (Bejder et al., 2019). In a laboratory setting, minimal changes in Hbmass, as low as 1 g.kg^-1^, can be accompanied by a significant change in aerobic capacity (Schmidt et al., 2010). To that extent we confirmed the previously reported link between absolute Hbmass and aerobic capacity in elite cyclists (Garvican et al., 2011;Hauser et al., 2017). It could therefore be argued that Hbmass would be a valid marker to complement the ABP analysis. The lack of relative influence of Hbmass (in g.kg^-1^ bodyweight) on time-trial performance may support the effect of a higher PV to reduce peripheral resistance to improve oxygen delivery to the muscle (Warburton et al., 2000;Mairbäurl, 2013). The relative weight of variations in single biomarkers in influencing the ABP markers should therefore be interpreted with care. This underlines the key role of ABP experts for a qualitative interpretation of suspicious profiles by accounting for and discriminating all possible confounders properly.

In our case we must underline that our cohort was composed exclusively of highly-trained elite cyclists and the first one collecting and interpreting monthly blood samples together with quantification of Hbmass and training load. Half of the participating athletes were part of a registered testing pool and subject to anti-doping testing and ABP profiling. Our findings may thus adequately reflect the situation found in an anti-doping context analyzing ABP profiles of elite athletes; and the variations observed in our cyclists are likely similar to those seen in other endurance sports (e.g., long distance running, cross-country skiing or biathlon). We must however acknowledge our small sample size limiting the power of our inferential analyses. In addition, the quantification of training load is notoriously difficult. Arguably our approach using TSS (combining intensity from power output and volume with training duration) was deemed the most pertinent when designing the study, with an interface routinely used by all our cyclists and their trainers. Characterizing objectively short periods of high acute training load before a blood test definitely remains challenging with the numerous training strategies possible. A simple declaration of high ATL (as a pretended alternative to blood withdrawal) may not be considered ultimately by an ABP expert as a unique pertinent explanation for a drop in [Hb]. Nevertheless, based on our findings we see a rationale for the inclusion of more complete information on training load on the days preceding an ABP sampling procedure.

In conclusion, we consider the ABP as a powerful tool for targeting anti-doping tests, and indirect detection of doping. Our study suggests that variations of acute training load (i.e. the 5 days before a sample is collected) may influence the ABP readings. Considering specific confounding factors (i.e. training load) is therefore certainly paramount in the qualitative assessment of variations observed in ABP profiles to adequately aim for cost-effective testing plans targeting the right athletes at the right moment.

## Data Availability

All data referred to in the manuscript may be requested to the corresponding author.

## Author Contributions

RF and MS conceived the project and obtained the project funding. TA and RF contributed to the collection of data. TA, RF and FCvR statistically analyzed the data. RF, TA, BK, FCvR and MS interpreted the data. TA wrote the first draft of the manuscript. All authors contributed to revising the manuscript and expressed their approval of the final submitted version.

## Acknowledgments

The authors wish to acknowledge WADA’s Science Department for the financial support of this study and all the participants for their participation.

## Supplementary files

**Figure 5.**
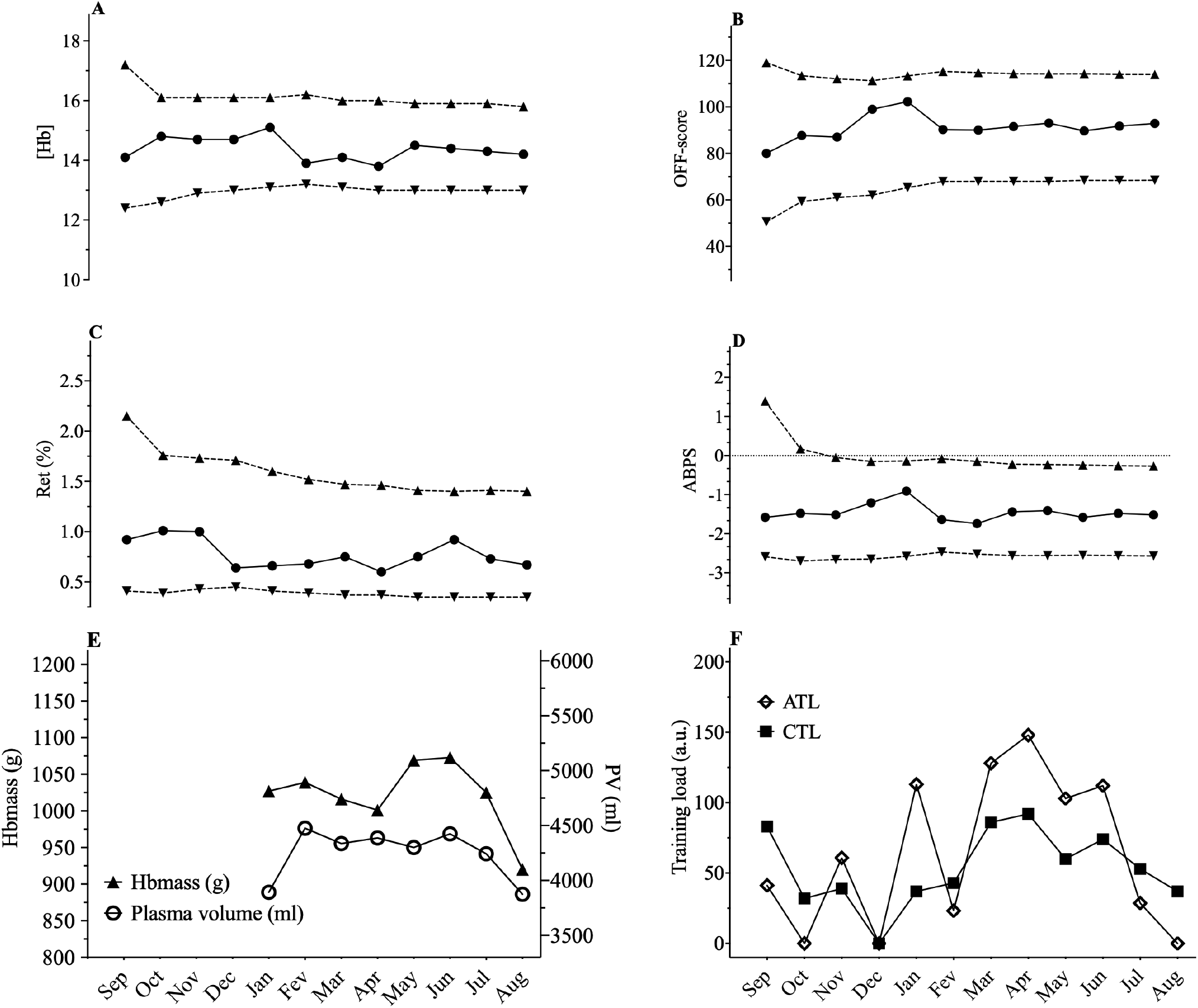
Representation of the Athlete Biological Passport (ABP) hematological profile for cyclist 1, with **(A)** [Hb]: hemoglobin concentration; **(B)** OFF-score, **(C)** Ret%: reticulocytes percentage; **(D)** ABPS: abnormal blood profile score; over the 12 months of the study design. Solid lines represents the athlete’s values, dotted line represents the upper limits and lower limits calculated by an adaptive Bayesian model (see methods section for details). **(E)** Acute training load (ATL) and chronic training load (CTL) represent the load respectively 5 and 42 days before sampling **(F)**. Triangles figure total hemoglobin mass (g) and circles represent plasma volume (mL) over the last eight months of the study.

**Figure 6.**
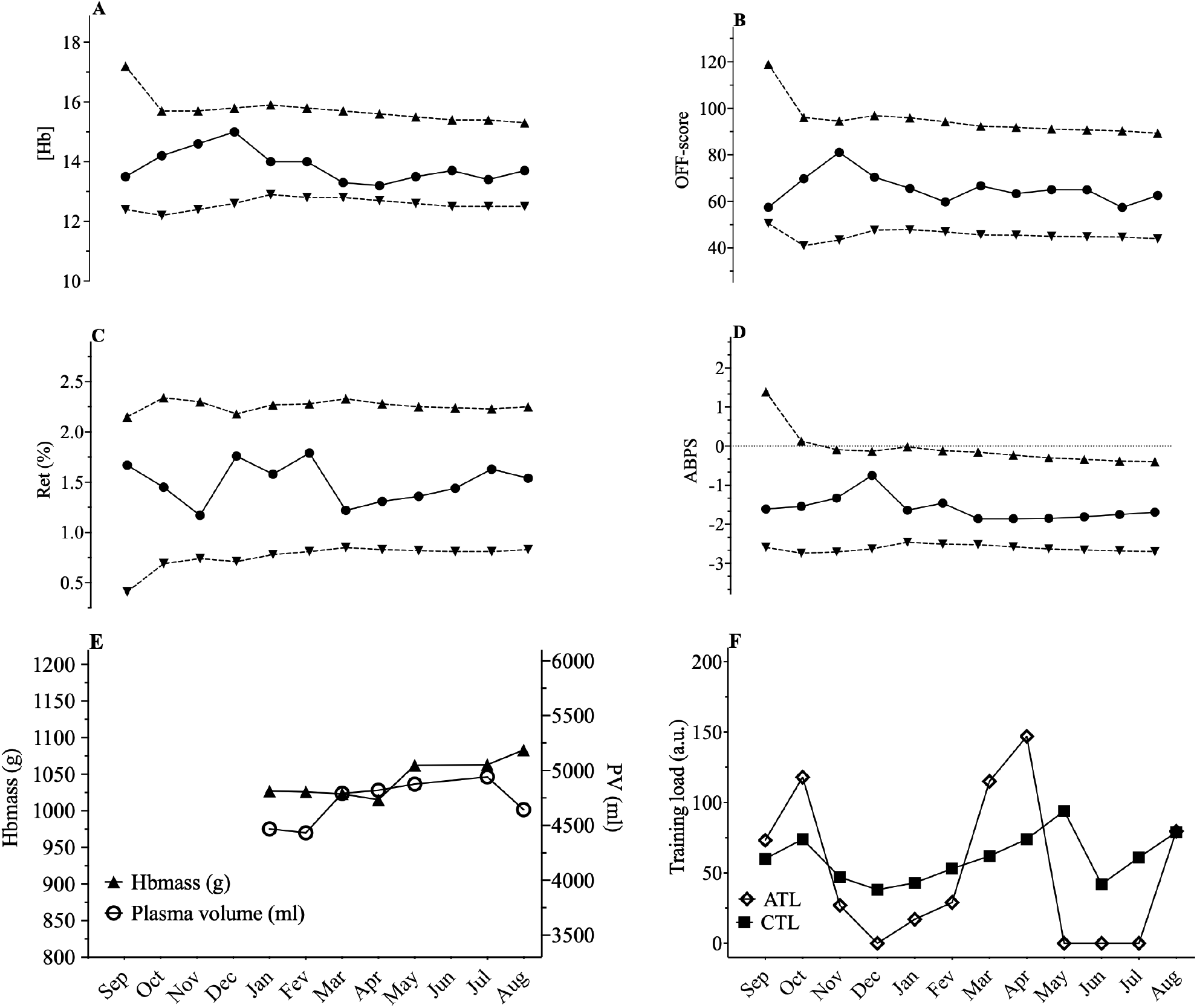
Representation of the Athlete Biological Passport (ABP) hematological profile for cyclist 3, with **(A)** [Hb]: hemoglobin concentration; **(B)** OFF-score, **(C)** Ret%: reticulocytes percentage; **(D)** ABPS: abnormal blood profile score; over the 12 months of the study design. Solid lines represents the athlete’s values, dotted line represents the upper limits and lower limits calculated by an adaptive Bayesian model (see methods section for details). **(E)** Acute training load (ATL) and chronic training load (CTL) represent the load respectively 5 and 42 days before sampling **(F)**. Triangles figure total hemoglobin mass (g) and circles represent plasma volume (mL) over the last eight months of the study.

**Figure 7.**
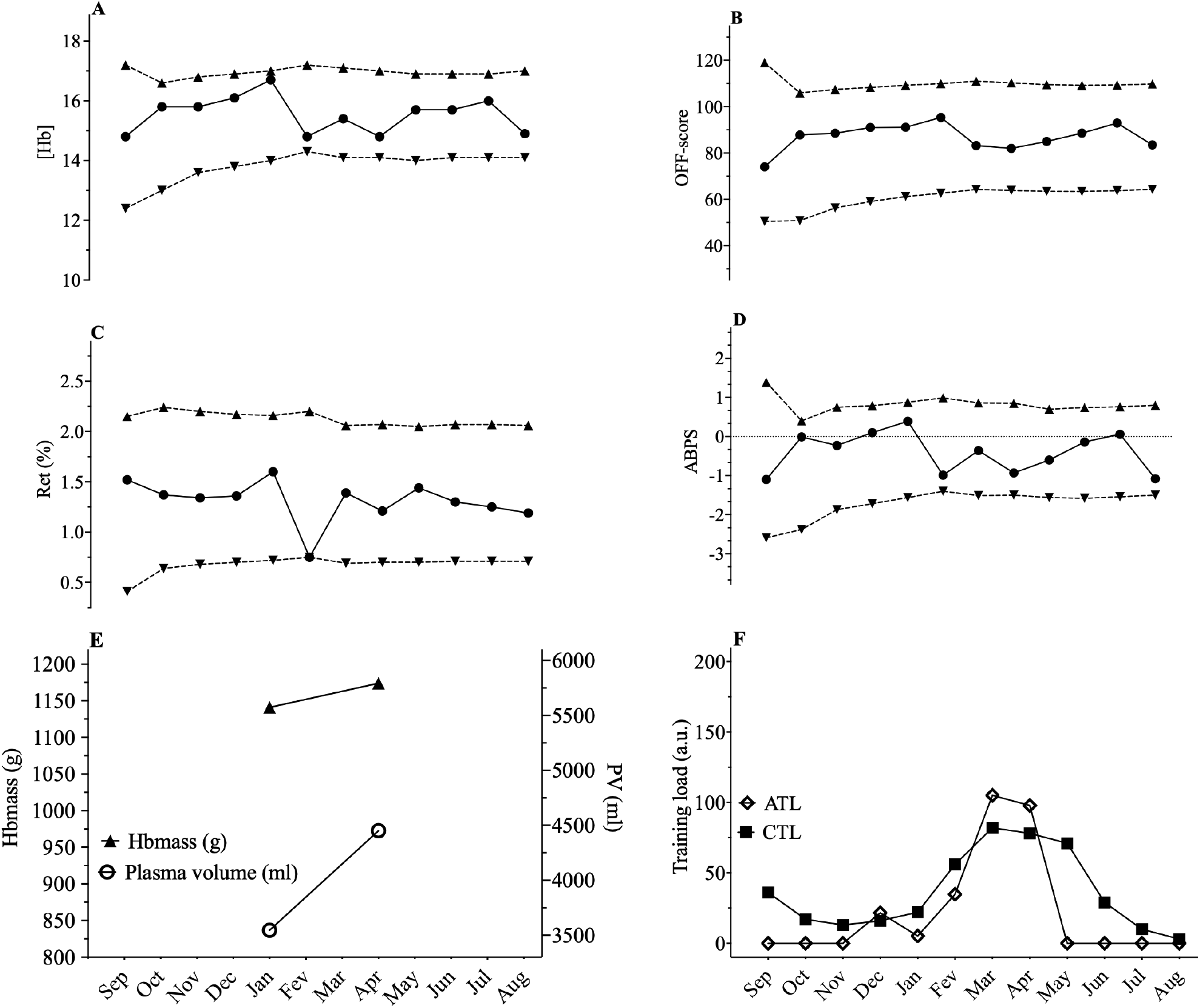
Representation of the Athlete Biological Passport (ABP) hematological profile for cyclist 4, with **(A)** [Hb]: hemoglobin concentration; **(B)** OFF-score, **(C)** Ret%: reticulocytes percentage; **(D)** ABPS: abnormal blood profile score; over the 12 months of the study design. Solid line represents the athlete’s values, dotted lines represents the upper limits and lower limits calculated by an adaptive Bayesian model (see methods section for details). **(E)** Acute training load (ATL) and chronic training load (CTL) represent the load respectively 5 and 42 days before sampling **(F)**. Triangles figure total hemoglobin mass (g) and circles represent plasma volume (mL) over the last eight months of the study.

**Figure 8.**
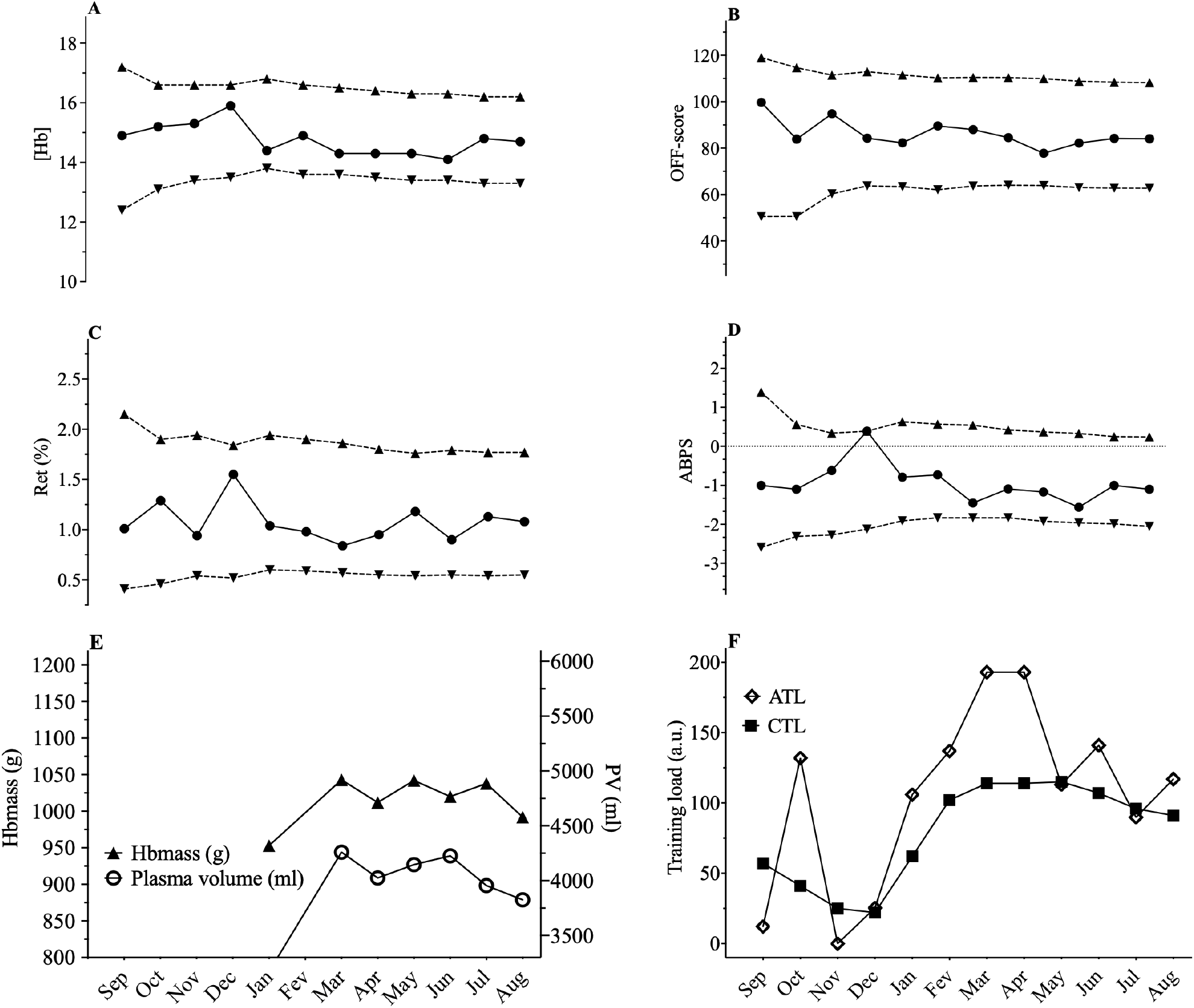
Representation of the Athlete Biological Passport (ABP) hematological profile for cyclist 5, with **(A)** [Hb]: hemoglobin concentration; **(B)** OFF-score, **(C)** Ret%: reticulocytes percentage; **(D)** ABPS: abnormal blood profile score; over the 12 months of the study design. Solid line represents the athlete’s values, dotted lines represents the upper limits and lower limits calculated by an adaptive Bayesian model (see methods section for details). **(E)** Acute training load (ATL) and chronic training load (CTL) represent the load respectively 5 and 42 days before sampling **(F)**. Triangles figure total hemoglobin mass (g) and circles represent plasma volume (mL) over the last eight months of the study.

**Figure 9.**
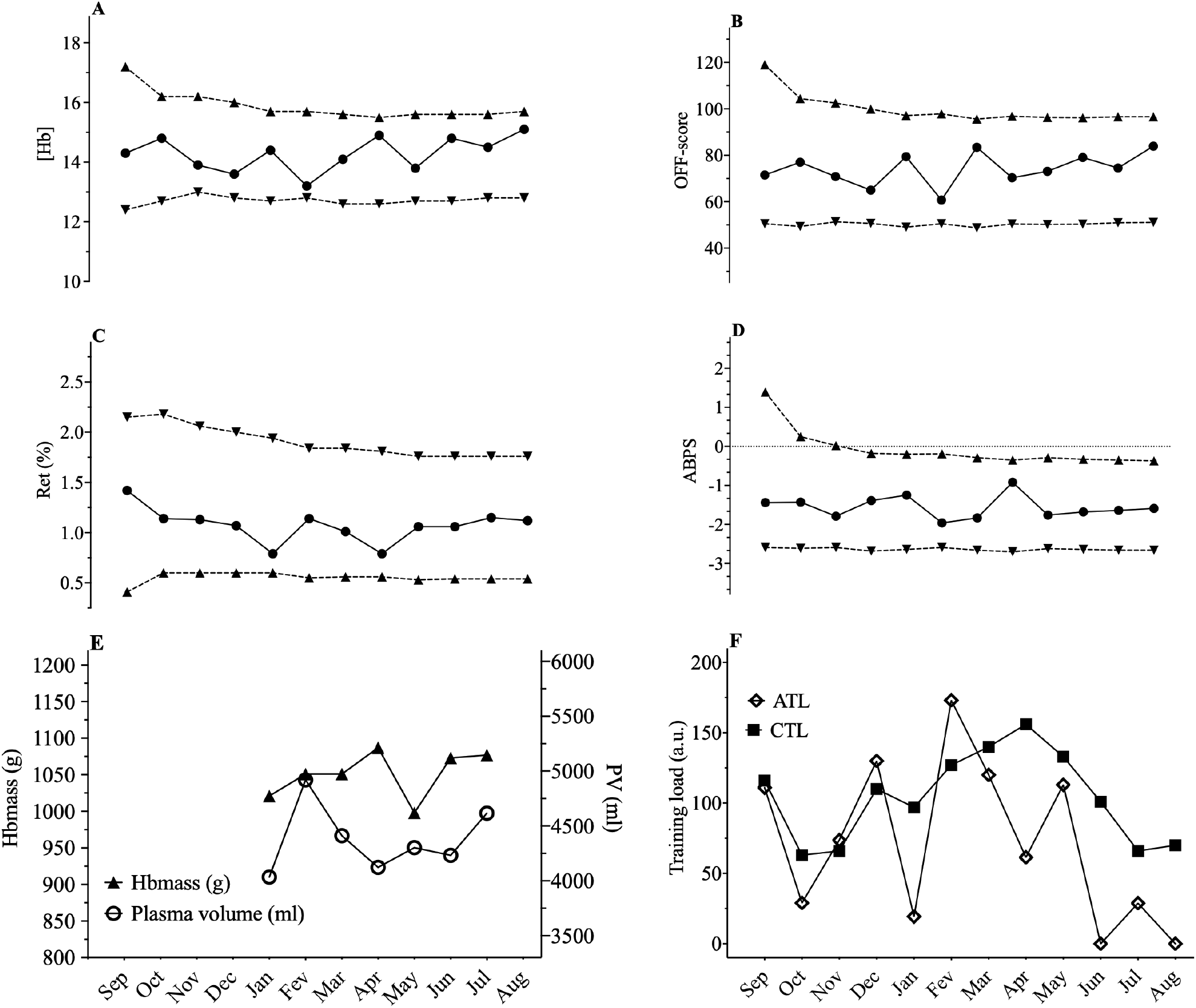
Representation of the Athlete Biological Passport (ABP) hematological profile for cyclist 8, with **(A)** [Hb]: hemoglobin concentration; **(B)** OFF-score, **(C)** Ret%: reticulocytes percentage; **(D)** ABPS: abnormal blood profile score; over the 12 months of the study design. Solid line represents the athlete’s values, dotted lines represents the upper limits and lower limits calculated by an adaptive Bayesian model (see methods section for details). **(E)** Acute training load (ATL) and chronic training load (CTL) represent the load respectively 5 and 42 days before sampling **(F)**. Triangles figure total hemoglobin mass (g) and circles represent plasma volume (mL) over the last eight months of the study.

**Figure 10.**
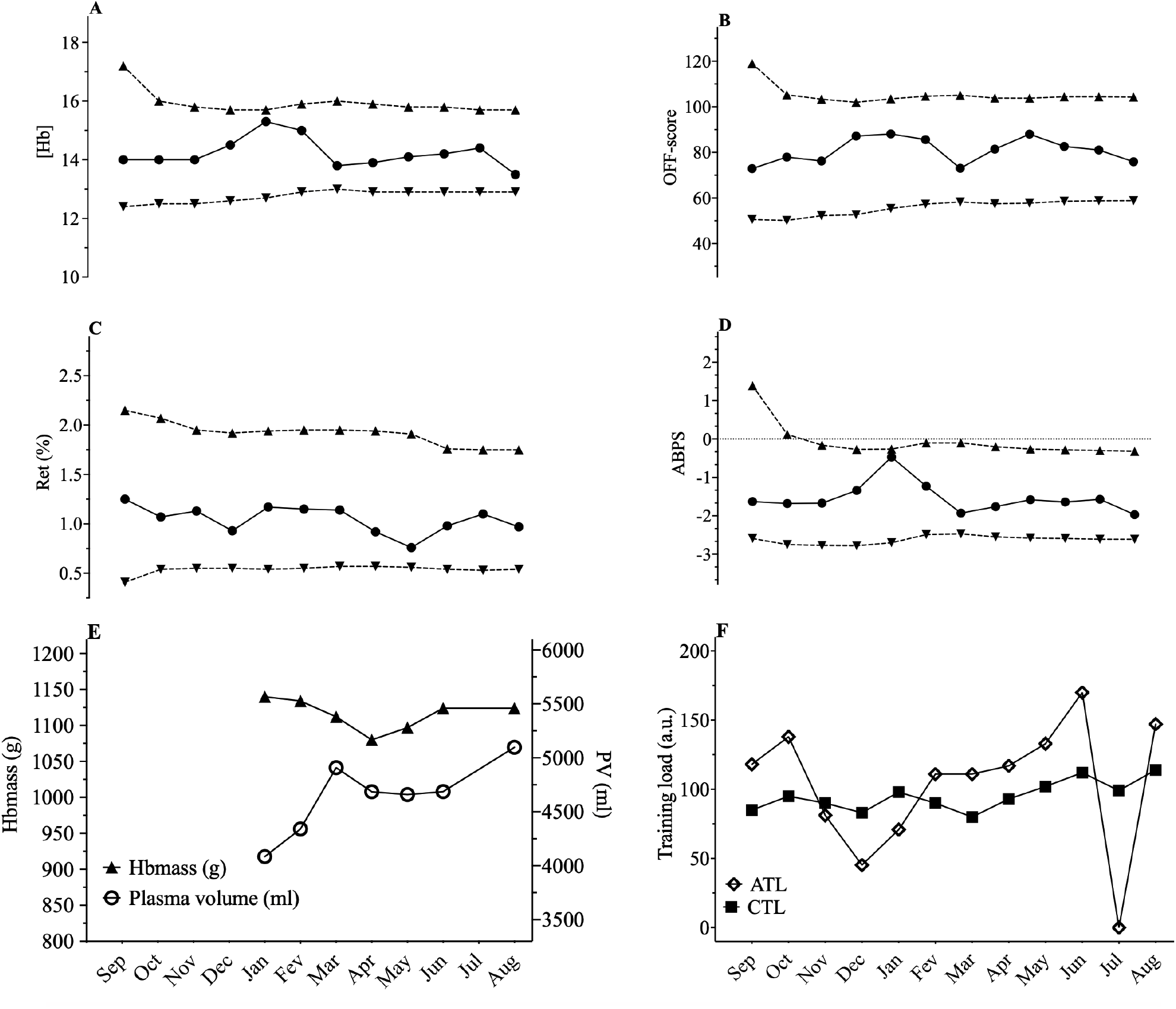
Representation of the Athlete Biological Passport (ABP) hematological profile for cyclist 9, with **(A)** [Hb]: hemoglobin concentration; **(B)** OFF-score, **(C)** Ret%: reticulocytes percentage; **(D)** ABPS: abnormal blood profile score; over the 12 months of the study design. Solid line represents the athlete’s values, dotted lines represents the upper limits and lower limits calculated by an adaptive Bayesian model (see methods section for details). **(E)** Acute training load (ATL) and chronic training load (CTL) represent the load respectively 5 and 42 days before sampling **(F)**. Triangles figure total hemoglobin mass (g) and circles represent plasma volume (mL) over the last eight months of the study.

**Figure 11.**
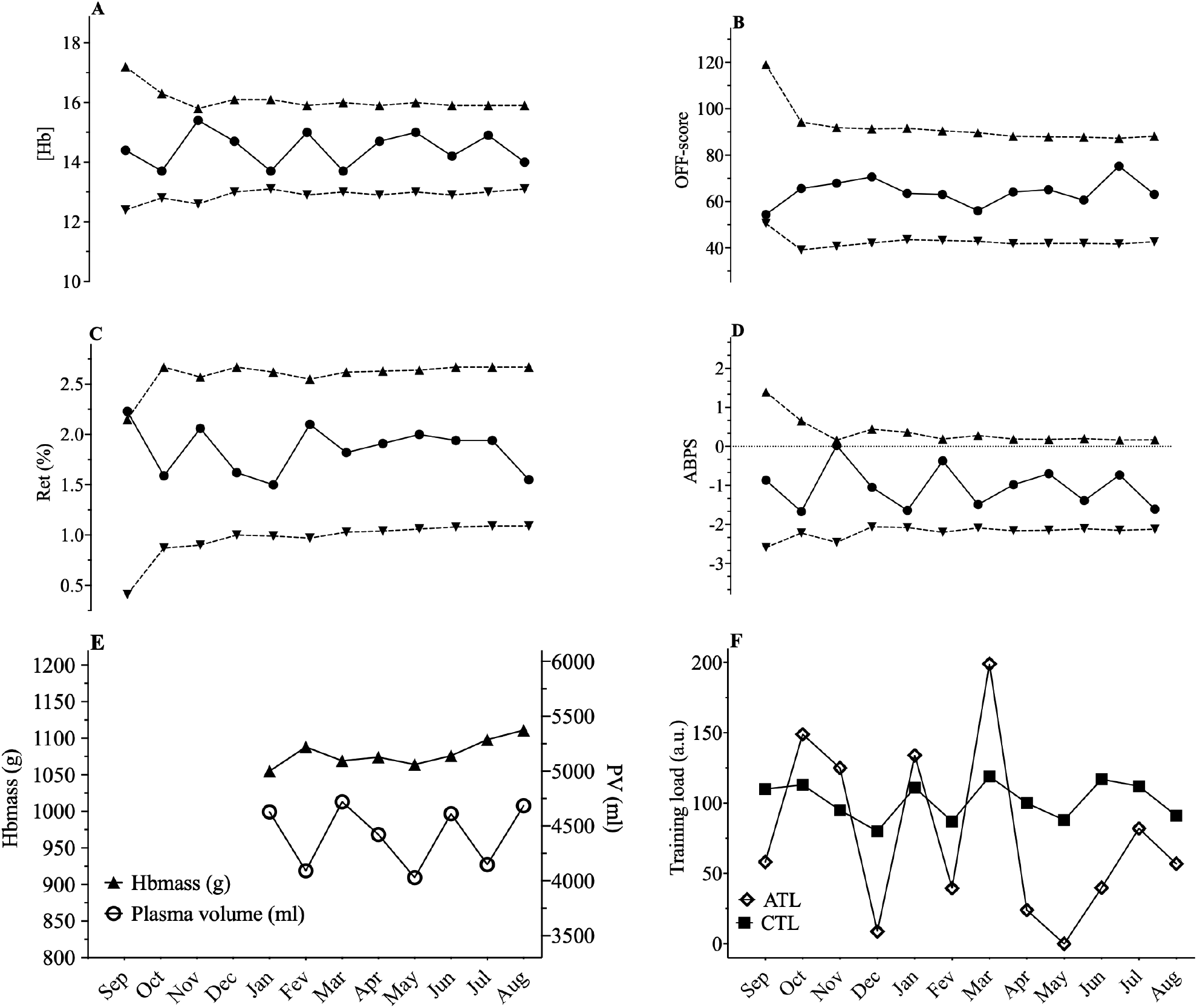
Representation of the Athlete Biological Passport (ABP) hematological profile for cyclist 10, with **(A)** [Hb]: hemoglobin concentration; **(B)** OFF-score, **(C)** Ret%: reticulocytes percentage; **(D)** ABPS: abnormal blood profile score; over the 12 months of the study design. Solid line represents the athlete’s values, dotted lines represents the upper limits and lower limits calculated by an adaptive Bayesian model (see methods section for details). **(E)** Acute training load (ATL) and chronic training load (CTL) represent the load respectively 5 and 42 days before sampling **(F)**. Triangles figure total hemoglobin mass (g) and circles represent plasma volume (mL) over the last eight months of the study.

